# Assessing knowledge and risk perception of mosquitoes and mosquito-borne viruses among medical students in Cyprus: A cross-sectional MosquitoWise study

**DOI:** 10.64898/2026.10.15.26365696

**Authors:** Abdalla Modar Dali, Anastasia Vujicic, Dania Gato, Ender Volkan

**Author notes:** Corresponding Author: Ender Volkan, University of Nicosia Medical School, 93 Agiou Nikolaou Street, Engomi, 2408 Nicosia, Cyprus. These authors contributed equally to the production of this research manuscript (Anastasia Vujicic, Dania Gato).

## Abstract

**Background:** Cyprus has transitioned into a high-alert zone for mosquito-borne viruses. This necessitates a proactive assessment of its future physicians. Following the 2022 re-establishment of *Aedes aegypti* in Larnaca and *Aedes albopictus* in Limassol, Cyprus, the island is now biologically primed for local outbreaks of dengue, Zika and chikungunya, alongside *Culex*-borne West Nile virus. This study assessed medical students’ knowledge and risk perception.

**Methods:** A cross-sectional online survey administered the validated MosquitoWise Health Belief Model instrument to undergraduate-entry and graduate-entry students at a university in Cyprus in mid-2026.

**Results:** Of 1023 students, 119 responded (11.6%), with pre-clinical students overrepresented (*p*<0.001). Mean knowledge was 5.43/9, stronger on mosquito biology (72.5%) than virus knowledge (54.2%). Median severity was 6.00/7 against susceptibility 4.25/7. Knowledge differed by study stage (*p*<0.001), correlated with severity (*r*_s_ = 0.399) but not susceptibility.

**Conclusions:** This study establishes a critical baseline for pre-clinical medical students in Cyprus. Overall, participants performed better on mosquito biology when compared to mosquito-borne viruses and how they spread. Our findings highlight the value of integrating theoretical microbiology with clinical practice in medical education to enhance physicians’ preparedness for the changing landscape of newly emerging infections in Cyprus and across Europe as a whole.

## Introduction

Human movement, urbanisation, and climate change are shaping the geographical distribution of disease-carrying vectors across the globe, such as the *Aedes* mosquitoes that transmit arboviral disease.^1^ While chikungunya and dengue have long been regarded in Europe as tropical illnesses found mostly in returning travellers,^2^ both are now being locally transmitted across the continent.^3^ Locally acquired *Aedes*-borne infections in Europe have risen steadily, with 43 of the 56 transmission events documented between 2007 and 2023 occurring in the final six years of that period.^4^

Cyprus’ unique position in the eastern Mediterranean makes the island highly susceptible to these emerging threats.^5^ Following the 2022 re-establishment of *Aedes aegypti* in the Larnaca region,^6^ alongside the first recorded detection of *Aedes albopictus* in Limassol in the same year,^7^ the island is now biologically primed for local outbreaks of *Aedes*-borne arboviruses,^1,8^ including dengue, Zika, and chikungunya. West Nile virus, transmitted principally by *Culex* species, has already been implicated in outbreaks in the region as well.^9^ In Cyprus itself, the 2019 outbreak was the island’s largest to date and included fatal cases,^5^ and transmission has resumed in 2026, with 14 cases being recorded, 13 of which had been locally acquired and one of them being fatal, according to Ministry of Health figures.^10^ This rapid shift highlights an urgent need for healthcare professionals equipped to handle such new threats.^11^

As the region confronts such challenges, the resilience of the healthcare system relies on the readiness of its future workforce.^12^ Medical students who will soon integrate into the Cypriot and other European healthcare systems must possess the clinical knowledge to identify these diseases and recognise the increasing risk of contraction in the general population. Establishing a baseline assessment of knowledge and risk perception (particularly in the early, pre-clinical years of education) is critical to identifying vulnerabilities before medical trainees enter hospital wards.^13,14^ Arboviral diseases have historically been presented within European clinical literature as a consideration in returning travellers rather than as a locally-acquired threat,^15^ which may leave trainees poorly calibrated to the possibility of domestic transmission.

The primary objective of the study was to evaluate the knowledge (KN) and risk perceptions of mosquitoes and mosquito-borne viruses among medical students. Using the MosquitoWise Survey, the study measured perceived susceptibility (SUS) and perceived severity (SEV). The secondary objective of the study was to assess how demographic subgroups (gender, programme of study, and stage of study) influence these results.

## Materials and methods

### Study design and setting

A cross-sectional observational study was conducted between April 17th and May 18th of 2026 at a medical school in Nicosia, Cyprus. An online survey was distributed via the European server instance of KoboToolbox (Kobo, Cambridge, MA, USA). The MosquitoWise Survey is based on the Health Belief Model (HBM) and was developed as part of the EU’s Horizon H2020 grant VEO (874735).^14^ The research was strictly observational. There were no clinical interventions done or biological samples collected.

### Participants and eligibility

The eligible population included 1023 medical students enrolled at a medical school in Nicosia, Cyprus aged 18 years or older and based in Cyprus, including 901 students enrolled in the Undergraduate-Entry Doctor of Medicine (MD) programme (6 years) and 122 students enrolled in the Graduate-Entry Doctor of Medicine (GEMD) programme (5 years). The programmes’ cohorts were consolidated into two stages: the pre-clinical basic sciences stage (Years 1 to 3 for both programmes) and the clinical sciences stage (Years 4 to 6 for the MD programme, and Years 4 to 5 for the GEMD programme) to allow for subgroup analysis by stage of study. The survey link was distributed to eligible participants via official student email addresses, with periodic reminders to encourage participation.

### Ethical considerations

All procedures contributing to this work comply with the ethical standards of the Declaration of Helsinki. The study received ethical approval from the Cyprus National Bioethics Committee (Application EEBK EΠ 2026.01.122). Electronic written informed consent was obtained on the first page of the online survey before any data were collected (the consent statement covers anonymous data collection, analysis, and retention for a period of five years). Participation was voluntary and all responses were anonymous. No personally identifying information was collected. Participants received no financial compensation for their participation.

### Study instrument and variables

Data was collected using the MosquitoWise Survey, a self-administered instrument based on the HBM and validated in general adult populations in the United Kingdom, Netherlands, and Spain.^14^ Because the published English version was designed for residents of the United Kingdom, the country and region of residency items were modified to fit Cyprus and its regions. A question covering the medical programme of study (MD or GEMD) and year of study was added. The validated HBM items were left unchanged. The outcome variables were the KN score and two HBM constructs, SUS and SEV.

The KN score was derived from four items assessing mosquito biting behaviour (KNbite), breeding sites (KNbreed), mosquito-borne viruses (KNvirus), and transmission routes (KNroute), scored using the same approach used by the instrument’s developers. ^16^ One point was awarded for each correctly selected response option, with no deductions for incorrectly selected options and no points awarded for “None of the above” or “I do not know” responses. Items were grouped into two subdomains: mosquito knowledge (KNbite, KNbreed; range 0 to 3 points) and mosquito-borne virus knowledge (KNvirus, KNroute; range 0 to 6 points), which were summed to yield a total score of 0 to 9, with higher scores indicating a greater level of knowledge.

SUS was calculated as the sum of four items (SUSmosq, SUSbite, SUSmbv, SUSres) and SEV as the sum of three items (SEVprobs, SEVdeadly, SEVqual), each rated on a seven-point Likert-type scale coded from 1 (“Strongly Disagree”) to 7 (“Strongly Agree”).^14,16^ No items required reverse coding. Total scores could range from 4 to 28 for SUS and 3 to 21 for SEV, with higher scores indicating greater perceived susceptibility and severity. Mean construct scores (possible range 1 to 7) were additionally calculated by dividing each sum by its number of items to allow for comparison with previously published European samples, consistent with the scoring methods used by the instrument’s developers.^16^ The two attention-check items (ControlA, ControlB) were used solely for data-quality control and were excluded from all scale scores.

The remaining constructs collected by the instrument (perceived barriers, benefits, self-efficacy, cues to action, prevention-measure use, and perceived responsibility) were not analysed in the present study. Independent grouping variables were gender, medical programme of study (MD or GEMD), and stage of study (pre-clinical or clinical).

### Bias and data quality control

The survey included two attention checks to identify satisficing responses:^17^ “ControlA” required the participant to choose the “Somewhat Agree” response, and “ControlB” required a “Disagree” response. Responses that failed either of these controls were excluded from the final dataset used for analysis. This exclusion criterion is stricter than the one applied in the original survey, which excluded participants only when both items were answered incorrectly;^16^ the stricter criterion allowed us to maximise confidence in response validity.

Out of 136 initial responses (gross response rate 136/1023, 13.3%), 17 were excluded (10 failed both control items and seven failed only one). The final sample contained 119 valid responses. All survey items were mandatory in KoboToolbox and only completed responses could be submitted. As a result, all 136 initial responses and 119 valid responses were complete. No missing data was present in any of the variables. No exclusion criteria based on completion time were applied.

Non-response bias was assessed by comparing the gender, medical programme of study, and stage of study of the sample population against official registrar data using Chi-square Goodness-of-fit testing. Due to the registrar’s binary (male / female) data, the seven respondents who selected “I prefer not to disclose” were excluded.

### Sample size and statistical analysis

The target sample size was determined on the basis of descriptive precision. A finite population correction was applied to the population of 1023 students, determining that 118 responses were required to estimate a population proportion with a margin of error of 8.5% at a 95% confidence level. The achieved sample of 119 valid responses satisfied this criterion.

Response rates, the sample size calculation and percentage-of-maximum scores were computed manually; all inferential analysis was done using jamovi (Version 2.7; The jamovi project, Sydney, Australia). Descriptive statistics (frequencies, percentages, means, standard deviations, medians, and interquartile ranges) were used to summarise knowledge scores and Likert-scale responses. Internal consistency was measured using Cronbach’s alpha across the nine scored components of the KN domain and the constituent items of the SUS and SEV domains respectively.

Outcome variables were compared across the two medical programmes of study using Independent Samples T-Tests. Homogeneity of variance was assessed via Levene’s test. Comparisons across the three gender categories were done using Welch’s One-Way ANOVA, accommodating more than two groups without assuming equal variances. Comparisons between the two stages of study used Welch’s procedure, equivalent for two groups to a Welch’s t-test. Bivariate associations between KN scores and the HBM constructs were evaluated using Spearman’s rank-order correlation coefficients, with 95% confidence intervals obtained by Fisher z transformation. The standardised mean difference for the primary two-group comparison is reported as Hedges’ g. Chi-square Goodness-of-fit tests were used to assess sample proportionality against expected frequencies derived from registrar data for gender, medical programme of study, and stage of study. Non-disclosed gender responses were excluded from the gender proportionality analysis to align with the registrar’s binary gender data, but were retained as a distinct category in all outcome comparisons by gender.

To assess the influence of the positive-only knowledge scoring approach, a sensitivity analysis was conducted in which one point was deducted for each incorrectly selected response option (floored at zero), and the primary analyses were repeated. Statistical significance was set at *p*<0.05. Given the exploratory nature of the secondary subgroup objectives, no formal correction for multiple comparisons was applied.

## Results

### Response rate and data quality

A total of 119 valid responses were used for the final analysis, yielding a net response rate of 119/1023 (11.6%). Response rates were closely comparable across the two medical programmes (MD: 105 of 901, 11.7% and GEMD: 14 of 122, 11.5%) but diverged sharply by stage of study, with pre-clinical students responding at more than twice the rate of clinical students (101 of 717, 14.1% vs 18 of 306, 5.9%). The analysed constructs demonstrated internal consistency within the study sample (KN α = 0.746 across nine components; SUS α = 0.768; SEV α = 0.803). Levene’s test showed homogeneity of variance across the two medical programmes for every measure (total KN *p*=0.86; mosquito subdomain *p*=0.20; virus subdomain *p*=0.89; SUS *p*=0.48; SEV *p*=0.35).

### Sample demographics and proportionality

Participant characteristics and sample proportionality are summarised in Table 1. Participants had a mean age of 21.5 years (SD = 3.0, range 18 to 37); as expected from the entry requirements, GEMD participants were older than MD participants. Most lived in urban areas and most had travelled outside their country of residency in the preceding two years.

**Table 1.** Participant characteristics and sample proportionality against the eligible student population (n=119)

**Panel A. Participant characteristics**
| Characteristic | Value |
| --- | --- |
| **Age (y)** |  |
| Mean (SD) | 21.5 (3.0) |
| Median | 21 |
| Range | 18–37 |
| MD, mean (SD) [range] | 20.9 (2.4) [18–32] |
| GEMD, mean (SD) [range] | 25.8 (3.9) [22–37] |
| **Gender, n (%)** |  |
| Woman | 66 (55.5) |
| Man | 46 (38.7) |
| Prefer not to disclose | 7 (5.9) |
| **Programme of study, n (%)** |  |
| MD | 105 (88.2) |
| GEMD | 14 (11.8) |
| **Stage of study, n (%)** |  |
| Pre-clinical | 101 (84.9) |
| Clinical | 18 (15.1) |
| **Year of study, n (%)** |  |
| MD year 1 | 27 (22.7) |
| MD year 2 | 19 (16.0) |
| MD year 3 | 43 (36.1) |
| MD year 4 | 13 (10.9) |
| MD year 5 | 2 (1.7) |
| MD year 6 | 1 (0.8) |
| GEMD year 1 | 5 (4.2) |
| GEMD year 2 | 5 (4.2) |
| GEMD year 3 | 2 (1.7) |
| GEMD year 4 | 1 (0.8) |
| GEMD year 5 | 1 (0.8) |
| **Area of residence, n (%)** |  |
| Urban | 96 (80.7) |
| Rural | 14 (11.8) |
| Do not know | 9 (7.6) |
| **Travel outside country of residency, preceding 2 y, n (%)** |  |
| Within Europe only | 33 (27.7) |
| Outside Europe only | 30 (25.2) |
| Both within and outside Europe | 43 (36.1) |
| No travel | 13 (10.9) |

**Panel B. Sample proportionality against registrar data (chi-square goodness-of-fit)**
| Characteristic | Category | Population, n (%) | Observed, n (%) | Expected, n (%) | $\chi^2$ (df) | *p* |
| --- | --- | --- | --- | --- | --- | --- |
| Gender<sup>a</sup> | Woman | 620 (60.6) | 66 (58.9) | 67.88 (60.6) | 0.132 (1) | NS |
|  | Man | 403 (39.4) | 46 (41.1) | 44.12 (39.4) |  |  |
| Programme | MD | 901 (88.1) | 105 (88.2) | 104.81 (88.1) | 0.003 (1) | NS |
|  | GEMD | 122 (11.9) | 14 (11.8) | 14.19 (11.9) |  |  |
| Stage | Pre-clinical | 717 (70.1) | 101 (84.9) | 83.40 (70.1) | 12.410 (1) | <0.001 |
|  | Clinical | 306 (29.9) | 18 (15.1) | 35.60 (29.9) |  |  |

*Note.* Bold denotes category headings; statistical symbols are italicised. Pre-clinical: years 1–3 of both programmes; clinical: years 4–6 (MD) and years 4–5 (GEMD). Eligible population N=1023. Panel A percentages are of n=119. Expected frequencies were derived from the university's registrar records (620 women, 403 men; 901 MD, 122 GEMD; 717 pre-clinical, 306 clinical). NS: not significant. There were no missing data for any variable. Percentages may not sum to 100 because of rounding.
<sup>a</sup> Participants who did not disclose a gender (n=7) were excluded from this comparison to match the registrar's binary records; percentages are of n=112. They were retained as a separate category in all outcome comparisons.

The sample did not differ detectably from population proportions by gender or by medical programme (Table 1). The seven participants who did not disclose a gender were excluded from the gender comparison to align with the registrar’s binary records.

Proportionality by stage of study, however, revealed a statistically significant non-response bias (Table 1): pre-clinical students were substantially overrepresented and clinical students underrepresented relative to the eligible population.

### Knowledge scores

KN scores by subgroup are presented in Table 2 and item-level responses in Table 3. The overall mean KN score was 5.43 (SD = 2.26) of a maximum of 9. Scores did not differ by medical programme or by gender, but clinical students scored substantially higher than pre-clinical students, a difference of 1.85 points (95% CI 0.92 to 2.78; Table 2).

**Table 2.** Knowledge scores by demographic subgroup (n=119)

A. By gender
| Knowledge measure (range) | All (n=119) | Woman (n=66) | Man (n=46) | Not disclosed (n=7) | Test statistic | *p* |
| --- | --- | --- | --- | --- | --- | --- |
| Total knowledge score (0–9) | 5.43 (2.26) | 5.29 (2.46) | 5.54 (2.04) | 6.00 (1.73) | *F*(2, 18.1)=0.519 | NS |

**Panel B. By programme of study**
| Knowledge measure (range) | All (n=119) | MD (n=105) | GEMD (n=14) | Difference (95% CI) | Test statistic | *p* | Hedges' *g* |
| --- | --- | --- | --- | --- | --- | --- | --- |
| Total knowledge score (0–9) | 5.43 (2.26) | 5.38 (2.25) | 5.79 (2.39) | 0.40 (–0.87 to 1.68) | *t*(117)=0.628 | NS | 0.18 |
| Mosquito subdomain (0–3) | 2.18 (1.01)<sup>a</sup> | 2.13 (1.02) | 2.50 (0.85) | 0.37 (–0.20 to 0.93) | *t*(117)=1.285 | NS | 0.36 |
| Virus subdomain (0–6) | 3.25 (1.74)<sup>b</sup> | 3.25 (1.73) | 3.29 (1.90) | 0.04 (–0.95 to 1.02) | *t*(117)=0.077 | NS | 0.02 |

**Panel C. By stage of study**
| Knowledge measure (range) | All (n=119) | Pre-clinical (n=101) | Clinical (n=18) | Difference (95% CI) | Test statistic | *p* | Hedges' *g* |
| --- | --- | --- | --- | --- | --- | --- | --- |
| Total knowledge score (0–9)<sup>c</sup> | 5.43 (2.26) | 5.15 (2.24) | 7.00 (1.68) | 1.85 (0.92 to 2.78) | *F*(1, 29.0)=16.589 | <0.001 | 0.85 |
| Mosquito subdomain (0–3) | 2.18 (1.01)<sup>a</sup> | 2.09 (1.04) | 2.67 (0.59) | 0.58 (0.23 to 0.93) | *F*(1, 38.7)=11.002 | 0.002 | 0.58 |
| Virus subdomain (0–6) | 3.25 (1.74)<sup>b</sup> | 3.06 (1.72) | 4.33 (1.46) | 1.27 (0.49 to 2.06) | *F*(1, 26.2)=11.046 | 0.003 | 0.75 |

*Note.* Bold denotes category headings; statistical symbols are italicised. Values are mean (SD) unless stated. Knowledge was scored positively, with one point per correctly selected option and no deductions for incorrect selections; total knowledge is the sum of the two subdomains. Gender was compared by Welch's one-way ANOVA at total-score level only, without mean differences or effect sizes. Programmes were compared by independent-samples *t*-test, with Levene's test confirming homogeneity of variance for all three measures. Stages were compared by Welch's procedure. Differences are expressed as GEMD – MD and clinical – pre-clinical. Hedges' *g* was calculated with the pooled SD. No correction for multiple comparisons was applied. NS: not significant. There were no missing data for any variable.
<sup>a</sup> 72.5% of the maximum attainable score. <sup>b</sup> 54.2% of the maximum attainable score.
<sup>c</sup> Sensitivity analysis applying penalty-based scoring (one point deducted per incorrectly selected option, floored at zero): overall 4.91 (SD 2.27); clinical 6.11 (SD 1.88) vs pre-clinical 4.69 (SD 2.27); difference 1.42 (95% CI 0.40 to 2.44); *F*(1, 26.8)=8.155; *p*=0.008; Hedges' *g*=0.63. Penalty-scored results were not disaggregated by gender or programme of study.

**Table 3.** Item-level knowledge responses (n=119)

| **Knowledge item** | **Participants, n** | **%** |
| --- | --- | --- |
| **Answered correctly: mosquito biology** |  |  |
| Rejected that mosquitoes bite only during daytime | 97 | 81.5 |
| Identified standing water in a gutter as a breeding site | 84 | 70.6 |
| Identified standing water in a bucket as a breeding site | 78 | 65.5 |
| **Correctly identified each mosquito-borne virus** |  |  |
| West Nile virus | 72 | 60.5 |
| Zika virus | 69 | 58.0 |
| Dengue virus | 66 | 55.5 |
| Chikungunya virus | 45 | 37.8 |
| **Correctly identified each transmission route** |  |  |
| Mosquito bite | 116 | 97.5 |
| Sexual contact | 19 | 16.0 |
| **Selected each incorrect option** |  |  |
| Food waste as a breeding site | 34 | 28.6 |
| Selected all three breeding-site options | 27 | 22.7 |
| Influenza virus | 8 | 6.7 |
| Measles virus | 4 | 3.4 |
| HIV | 3 | 2.5 |
| Touching an infected person | 6 | 5.0 |
| Airborne transmission | 5 | 4.2 |
| Touching an infected animal | 2 | 1.7 |

*Note.* Bold denotes category headings; statistical symbols are italicised. Each row gives the number of participants who selected that response, as a percentage of the full sample. Items were multiple-select, so rows are not mutually exclusive and percentages do not sum to 100. The 27 participants who selected all three breeding-site options are a subset of the 34 who selected food waste. Incorrect options were recorded but not scored. There were no missing data for any variable.

Knowledge was unevenly distributed across subdomains. Participants performed better on mosquito biology (72.5% of maximum) than on mosquito-borne virus knowledge (54.2%), and both subdomains differed significantly by stage of study (Table 2). At item level (Table 3), most participants rejected the assertion that mosquitoes bite only during daytime and identified the mosquito bite as a transmission route, but identification of specific arboviruses was weaker, with chikungunya the least frequently identified of the four assessed (45/119, 37.8%). 19/119 (16.0%) recognised sexual contact as a possible transmission route.

### Perceived susceptibility and severity

The sample yielded a median SUS score of 4.25 out of 7 (IQR = 1.88, range 1.50 to 6.75), corresponding to a sum score of 17 out of 28. The median SEV score was 6.00 out of 7 (IQR = 1.00, range 3.33 to 7.00), corresponding to a sum score of 18 out of 21.

Subgroup analyses showed that these risk perceptions remained consistent regardless of the academic programme. No statistically significant differences were observed between the MD and GEMD cohorts in either SUS, *t*(117) = 1.209, *p*=0.23, or SEV, *t*(117) = 0.819, *p*=0.41. Furthermore, Welch’s ANOVAs confirmed that risk perception constructs did not differ significantly by stage of study for either SUS, *F*(1, 27.0) = 0.145, *p*=0.71, or SEV, *F*(1, 24.8) = 2.335, *p*=0.14. Risk perceptions likewise did not differ significantly across gender for either SUS, *F*(2, 17.8) = 0.808, *p*=0.46, or SEV, *F*(2, 17.2) = 2.753, *p*=0.09.

### Bivariate correlations

Spearman’s rank-order correlations were computed to assess the relationship between foundational knowledge and the HBM constructs. The analysis revealed a positive correlation between participants’ KN scores and their SEV scores (*r*_s_ = 0.399, 95% CI 0.24 to 0.54, *p*<0.001). Conversely, KN did not correlate detectably with SUS (*r*_s_ = −0.082, 95% CI −0.26 to 0.10, *p*=0.37). The correlation between SUS and SEV was likewise not statistically significant (*r*_s_ = 0.136, 95% CI −0.05 to 0.31, *p*=0.14).

### Sensitivity analysis

A sensitivity analysis applying a penalty-based scoring rule, deducting one point for each incorrectly selected response option, lowered the mean KN score but did not alter any substantive finding (Table 2). KN continued to correlate with SEV, *r*_s_ = 0.428, *p*<0.001, while showing no detectable correlation with SUS, *r*_s_ = −0.097, *p*=0.30.

## Discussion

This cross-sectional study established a baseline assessment of medical students’ knowledge and risk perceptions of mosquito-borne viruses in Cyprus, where West Nile virus already circulates and vectors for dengue, chikungunya, and Zika (*A. aegypti* and *A. albopictus*) are now present.^5^ The main knowledge findings show that participants performed better on mosquito biology (72.5%) than on mosquito-borne viruses and how they spread (54.2%). Participants regarded mosquito-borne illness as clinically serious while remaining unsure about their own susceptibility, and KN was associated with SEV but not with SUS.

The significant non-response bias skewed the sample heavily toward pre-clinical students, so this study effectively captures trainees across the pre-clinical years, the largest single group being in the final year before the clinical stage. Identifying these vulnerabilities early matters because the resilience of the Cypriot and broader European healthcare systems relies on physicians equipped to recognise the presentation and risks of mosquito-borne viruses.^11^

Analysing the participants’ perceptions through the validated HBM domains of the MosquitoWise Survey reveals a marked asymmetry in risk appraisal.^14^ The median SEV score corresponded exactly to the “Agree” anchor, while the median SUS score sat only marginally above the neutral midpoint. Students therefore regard mosquito-borne illness as clinically serious while remaining undecided as to whether it is likely to occur in their own setting. If risk perception remains misaligned with current realities, potentially stemming from the historical view of Europe as a low-risk region for tropical diseases,^2^ future physicians may fail to prioritise or recommend essential preventative practices.^11^ The framing of arboviral disease as a consideration in the returning traveller rather than a locally acquired threat, long embedded in European clinical literature,^15^ is now obsolete.

This asymmetry is reinforced by the correlational findings. KN was positively associated with SEV but showed no detectable association with SUS, and the two constructs were unrelated to one another. Factual knowledge about mosquito-borne viruses therefore appears dissociable from students’ sense of local risk. Students learn what these diseases are and what they do, but not that they now occur here in Europe. Future medical curricula must target these specific psychological constructs to help convert theoretical microbiology knowledge into an accurate interpretation of clinical severity and susceptibility.^11^

The subdomain analysis locates the deficit more precisely. Participants performed well on vector biology but less well on virus-specific knowledge. Chikungunya was the least frequently identified of the four arboviruses assessed, despite being transmitted by *A. albopictus*, the species with established presence in Limassol.^7^ The arbovirus posing one of the most immediate local transmission risks was the one participants were least able to name. Few participants recognised sexual contact as a transmission route, a consequential gap given its established role in Zika virus transmission and its implications for returning travellers, even though its contribution to overall transmission is small.^18^ The epidemiological weight of this route remains debated: viral RNA persists in semen for months, but infectious virus is recovered only briefly, and estimates of the share of infections attributable to sexual transmission remains uncertain.^18^

### Limitations

The study is subject to several methodological limitations. Primarily, the net response rate of 119/1023 (11.6%) restricts generalisability, permitting population estimates only within a margin of error of approximately 8.5% at a 95% confidence level, and the significant non-response bias by stage of study (Table 1) reflects clinical students under-participating, potentially because of survey fatigue and time constraints during rotations. The results function strictly as a baseline for pre-clinical students and should not be generalised to clinical students. Participants were also drawn from a single medical school, and the findings may not reflect students at other institutions in Cyprus or elsewhere in Europe, where curricular coverage of vector-borne disease may differ.

Additionally, because data was gathered via the self-reported online MosquitoWise Survey, responses are subject to social desirability bias, particularly on the agreement-scaled risk perception items.^19^ Recall bias applies to the travel history item, which required reporting holidays taken over the preceding two years.

The uneven distribution of participants across subgroups (Table 1) limited statistical power for the secondary comparisons. The non-significant findings for risk perception should therefore be interpreted as an absence of detected difference rather than as evidence of equivalence.

Analyses were restricted to the pre-specified subgroups of gender, medical programme, and stage of study. Although participant age was recorded, it was not examined as an independent predictor, as age is substantially collinear with both programme and stage of study, and the sample size did not support additional stratified comparisons.

Knowledge was scored positively, without deductions for incorrectly selected response options, to preserve comparability with previous studies employing the MosquitoWise Survey.^14,16^ This may inflate scores among participants who select indiscriminately: more than a quarter of participants selected the breeding-site distractor (Table 3). Over-selection was, however, concentrated on that single item, with every other incorrect option selected by fewer than one in ten participants, suggesting a specific misconception regarding food waste as a breeding substrate rather than generalised inattentive responding. A sensitivity analysis applying a penalty-based rule confirmed that no substantive conclusion depended on the scoring approach.

Finally, while the MosquitoWise Survey was developed and validated in general adult populations rather than in medical students, all three measures showed acceptable internal consistency (Cronbach’s alpha 0.746 to 0.803). Given that the knowledge items are each a factual question scored against correct answers, they do not depend on a factor structure.

## Conclusions

Among medical students surveyed at this university in Cyprus, awareness of mosquito-borne viruses varied, with chikungunya being the least frequently named arbovirus. This is significant considering the recent establishment of the chikungunya vector *Aedes albopictus* in Cyprus and its broader distribution in Europe with the ability to spread various pathological viruses. Students judged mosquito-borne illness to be serious while remaining unsure about their own exposure, and KN correlated positively with SEV while showing no detectable association with SUS. Learning what these diseases do is evidently separable from recognising that they now occur in Cyprus and Europe as a whole. Although single-institution findings still warrant further investigation in other education settings, they highlight the necessity of medical curricula to evolve and strengthen the integration of theoretical microbiology with the clinical reality of locally acquired arboviral infections.

## Authors’ contributions

A.M.D.: Methodology, Validation, Formal Analysis, Investigation, Resources, Data Curation, Writing – original draft, Writing – review & editing, Visualization, Project Administration. A.V.: Conceptualization, Methodology, Formal Analysis, Investigation, Writing – original draft, Writing – review & editing. D.G.: Conceptualization, Methodology, Investigation, Writing – original draft, Writing – review & editing. E.V.: Methodology, Validation, Investigation, Resources, Writing – review & editing, Visualization, Supervision, Project Administration. All authors read and approved the final manuscript.

## Acknowledgements

The authors used standard AI assistants (Claude, Anthropic and ChatGPT, OpenAI) to double-check the manuscript against the journal’s manuscript guidelines, suggest formatting edits, and cross-check the tables for consistency with the main text. The authors reviewed all suggestions before final implementation and take full responsibility for the content.

## Ethical approval

The study was approved by the Cyprus National Bioethics Committee (application EEBK EΠ 2026.01.122). All participants provided electronic informed consent before data collection.

## Data availability

The underlying data cannot be shared publicly because participant consent covered only anonymous data collection, analysis, and retention of the data by the research team for five years. The anonymised dataset may be provided on reasonable request to the corresponding author for the journal’s internal review and validation purposes.

